# Informing behaviour change intervention design using systematic review with Bayesian meta-analysis: physical activity in heart failure

**DOI:** 10.1101/2021.09.05.21262643

**Authors:** A. Amirova, L. Taylor, B. Volkmer, N. Ahmed, A.M. Chater, T. Fteropoulli

## Abstract

Embracing the Bayesian approach, we aimed to synthesise evidence regarding barriers and enablers to physical activity in heart failure (HF) in a way that can inform behaviour change intervention development. This approach helps in estimating and quantifying the uncertainty in the evidence and facilitates the synthesis of qualitative and quantitative studies. Qualitative and observational studies investigating barriers and enablers to physical activity in adults diagnosed with HF were included in this systematic review with a Bayesian meta-analysis. Qualitative evidence was annotated using the Theoretical Domains Framework and represented as a prior distribution using an expert elicitation task. The maximum a posteriori probability (*MAP*) was calculated as a summary statistic for the probability distribution for the *log OR* value estimating the relationship between physical activity and each determinant, according to qualitative evidence alone, quantitative evidence, and qualitative and quantitative evidence combined. The dispersion in the probability distribution for log OR associated with each barrier or enabler was used to evaluate the level of uncertainty in the evidence. Wide, medium, and narrow dispersion (SD) corresponded to high, moderate, and low uncertainty in the evidence, respectively. Evidence from three qualitative and 16 (*N* = 2739) quantitative studies was synthesised. High pro-b-type natriuretic peptide, pro-BNP (*MAP* value for *log OR* = -1.16; 95% *CrI*: [-1.21; -1.11]) and self-reported symptoms (*MAP* for *log OR* = 0.48; 95% *CrI*: [0.40; 0.55]) were suggested as barriers to physical activity with narrow distribution dispersion (*SD* = 0.18 and 0.19, respectively). Modifiable barriers were symptom distress (*MAP* for *log OR* = -0.46; 95% *CrI*: [-0.68; -0.24]), and negative attitude (*MAP* for *log OR* = -0.40; 95% *CrI*: [-0.49; -0.31]), *SD* = 0.36 and 0.26, respectively. Modifiable enablers were social support (*MAP* for *log OR* = 0.56; 95% *CrI*: [0.48; 0.63]), self-efficacy (*MAP* for *log OR* = 0.43; 95% *CrI*: [0.32; 0.54]), positive physical activity attitude (*MAP* for *log OR* = 0.92; 95% *CrI*: [0.77; 1.06]), *SD* = 0.26, 0.37, and 0.36, respectively. This work extends the limited research on the modifiable barriers and enablers for physical activity by individuals living with HF.

## Introduction

Heart Failure (HF) is a complex clinical syndrome of symptoms that suggest reduced efficiency with which the heart pumps blood around the body (National Institute for Healthcare and Excellence, 2018). It is a prevalent condition worldwide, affecting 2% of the general adult population (Groenewegen, Rutten, Mosterd, & Hoes, 2020). The condition affects older adults, with the prevalence rising to 6.6% among individuals aged 65 and over and to 13.5% among individuals aged 80 and over (Mozaffarian et al., 2015). HF is a debilitating condition characterised by symptoms of peripheral water retention (i.e., oedema), breathlessness, and fatigue. Despite advances in medical treatment, HF is a major cause of morbidity and mortality (Bragazzi et al., 2021).

Physical activity is associated with improved quality of life (Davies et al., 2010; Lewinter et al., 2015; Sagar et al., 2015; Taylor et al., 2019), reduced hospitalisation (Sagar et al., 2015) and increased longevity (Belardinelli, Georgiou, Cianci, & Purcaro, 2012; ExTraMATCH Collaborative, 2004) in individuals living with HF. Therefore, regular physical activity is a key component of recommended treatment (Ponikowski et al., 2016). While the minimal clinically important difference in physical activity levels in HF is not known (Dibben et al., 2018; Shoemaker, Curtis, Vangsnes, & Dickinson, 2013), the recommendation for older adults, in general, is to perform a minimum of 75–150 minutes per week of moderate to vigorous intensity aerobic physical activity; and engage in functional balance and muscle strength training at a moderate intensity at least three and two days a week, respectively (WHO, 2020).

A structured form of physical activity, exercise, is included in cardiac rehabilitation (CR) and is offered to newly diagnosed HF patients (National Institute for Healthcare and Excellence, 2018). However, uptake of CR programmes is less than 1% among individuals diagnosed with HF (Doherty & Harrison, 2017). Levels of everyday physical activity in HF are also low (Jaarsma et al., 2013; O’Donnell et al., 2020), partially due to the many challenges individuals with HF face in initiating and maintaining a physically active lifestyle, as proposed by the European Society of Cardiology (Conraads et al., 2012). Understanding how best to improve physical activity in individuals living with HF is warranted.

Interventions informed by a behaviour change theory are potentially promising in achieving physical activity improvements in HF (Tierney et al., 2011). A recent meta-analysis found that interventions integrating an exercise programme with behaviour change theory and interventions delivered by a physiotherapist are efficacious in obtaining short-term improvements in physical activity levels among individuals living with HF (Amirova, Fteropoulli, Williams, & Haddad, 2021). However, the effect of these interventions varied considerably. In addition, the extent to which a theory informed interventions was limited (Amirova, Fteropoulli, Williams, & Haddad, 2021). Research on the barriers and enablers influencing physical activity is needed. This may inform the choice of behaviour change theory and intervention development.

Guidelines for developing behaviour change interventions recognise that the modifiable and contextual barriers and enablers need to be systematically identified and described to inform intervention design (Araújo-Soares, Hankonen, Presseau, Rodrigues, & Sniehotta, 2019; Hagger, Cameron, Hamilton, Hankonen, & Lintunen, 2020; Kok et al., 2016; Michie, van Stralen, & West, 2011; O’Cathain, Croot, Duncan, et al., 2019). Knowledge about relevant determinants increases the intervention’s chances to be effective and conserves research effort and resources (Craig et al., 2008; O’Cathain, Croot, Sworn, et al., 2019). Systematically identified evidence concerning modifiable and contextual barriers and enablers can guide theory choice and therefore inform behaviour change intervention design.

However, the factors influencing physical activity participation in individuals living with HF are not well understood. A systematic review of qualitative studies found a lack of research on individual accounts of barriers and enablers to physical activity in individuals living with HF (Tierney et al., 2011). The review reported sparse summaries about physical activity extracted from the studies that elucidated beliefs and personal accounts of living with HF in general, including physical activity only as one of many themes (Tierney et al., 2011). The following enablers: knowledge of risks and benefits associated with physical activity (e.g., reduced mortality and morbidity, and improved quality of life); confidence in one’s ability to engage in physical activity; anticipated outcomes of physical activity; and social support, as well as a barrier such as weather, were previously identified in a systematic narrative review (Tierney et al., 2011). However, these barriers and enablers have not been confirmed in quantitative studies. The review highlighted the need to explore further what influences physical activity in HF (Tierney et al., 2011).

A recent paper called for the adoption of Bayesian statistics in Health Psychology research (Beard & West, 2017; Depaoli et al., 2017; Hamilton, Marques, & Johnson, 2017; Heino, Vuorre, & Hankonen, 2018). This approach may also be useful in understanding the contextual and modifiable determinants influencing physical activity in HF. The Bayesian approach views evidence synthesis as a decision-making process (Roberts, Dixon-Woods, Fitzpatrick, Abrams, & Jones, 2002); new evidence is considered in light of existing evidence, beliefs, and practices. Beliefs are often presented in the form of qualitative research. Qualitative research is readily available from research studies on health and health management; however, its findings are not utilised in healthcare decision-making and policy development (Roberts et al., 2002). Qualitative research provides rich data, but the required formal systematic evaluation impedes the inclusion of qualitative evidence in decision-making and policy development (Roberts et al., 2002). This makes it difficult for qualitative evidence to inform policy-making (Roberts et al., 2002). It is also recommended to account for stakeholders’ needs – the needs of those living with HF in this instance – in research concerning intervention development (Craig et al., 2008). While Bayesian methods provide an opportunity to incorporate qualitative evidence in decisions about health management (Spiegelhalter, Abrams, & Myles, 2003). Therefore, Bayesian methods are useful when evidence from diverse sources needs to be synthesised.

However, to perform Bayesian synthesis, qualitative research should be formally and systematically catalogued before it can be integrated with quantitative findings, which is often not straightforward (Roberts et al., 2002). The Theoretical Domains Framework (Cane, O’Connor, & Michie, 2012) is a tool developed through an international collaborative effort that systematically describes domains and constructs that may influence behaviour under investigation. In the current study, the identified physical activity barriers and enablers in HF were categorised in accordance with the TDF. In addition, the COM-B model, developed from a systematic synthesis of behaviour change frameworks (Michie et al., 2011), was used to inform future behaviour change interventions targeting physical activity in HF. In particular, following the consensus on the link between barriers and enablers, the intervention’s proposed mechanisms and strategies (Connell et al., 2019), and several behaviour change techniques (BCTTv1: (Michie et al., 2013) that are likely to enhance the identified relevant enablers or reduce the barriers were proposed.

### Objectives

The present review with meta-analysis aims to systematically integrate qualitative and quantitative evidence on the clinical, environmental, and psychosocial barriers and enablers influencing physical activity in those living with HF. The secondary aim, which is a response to the recent call to embrace the Bayesian approach (Beard & West, 2017; Depaoli et al., 2017; Hamilton et al., 2017; Heino et al., 2018), is to apply the Bayesian approach in synthesising evidence regarding barriers and enablers to physical activity in HF in a way that can inform behaviour change intervention development.

## Method

The systematic review with meta-analysis was implemented adhering to guidance on conducting systematic reviews and meta-analyses of observational studies of aetiology, COSMOS-E (Dekkers et al., 2019). The review is reported following PRISMA 2020 guidelines (Page et al., 2021). The review’s protocol was registered on PROSPERO: CRD42021232048.

### Eligibility criteria

Qualitative and observational studies investigating any clinical, environmental, social, or psychological barriers and enablers to physical activity in adults diagnosed with HF were included in this review (supplement 1). Physical activity was defined as any bodily movement that requires metabolic energy expenditure (WHO, 2010), of any mode (e.g., walking); any intensity (e.g. moderate to vigorous); in any setting (as exercise prescription or otherwise). For practical reasons, the search results were further restricted to peer-reviewed articles in English.

### Information sources

A total of 14 online databases were searched from inception to 05 January 2020 (Embase, Global Health, HMIC Health Management Information Consortium, MEDLINE; PsychINFO; CINAHL; Health policy reference centre; PsychARITCLES; PubMed; The Cochrane Library; Academic search complete, Pedro). The reference lists of the obtained articles included at full-text screening were hand searched for relevant studies meeting the inclusion criteria. In addition, ClinicalTrial.gov was searched for observational studies but yielded no results.

### Search strategy

The MeSH terms and keywords describing the Population of interest (i.e. HF and nine synonyms combined using a Boolean operator ‘OR’) and Outcome of interest (i.e. physical activity and 21 synonyms combined using a Boolean operator ‘OR’) were combined using a Boolean operator ‘AND’ (supplement 2). The initial search yielded 11,678 hits.

### Selection process

Two reviewers (AA and LT) independently screened titles and abstracts and selected articles meeting the criteria for full-text screening in Rayyan. Qualitative studies meeting the eligibility criteria informed the prior elicitation task (i.e., appraisal by a panel of experts). Quantitative studies were included in the frequentist meta-analysis.

### Data collection process

Two reviewers (AA and LT) independently extracted relevant data items from the reports of the included studies.

### Data items

The Strengthening the Reporting of Observational Studies in Epidemiology items, STROBE (von Elm et al., 2007) were utilised to design the data extraction form (supplement 3).

### Study risk of bias assessment

Two reviewers (AA and LT) independently assessed the study-level risk of bias present in the included quantitative studies. The following sources of bias were considered: selective reporting, participant selection, missing data (including non-respondents and dropouts), confounding (measured and unmeasured confounds; time-varying confounds in cohort studies), and outcome definition and measurement (i.e. information bias) (Dekkers et al., 2019). Due to the lack of a comprehensive risk of bias tool designed specifically for observational studies (Page, McKenzie, & Higgins, 2018), three instruments were used jointly: the Appraisal tool for Cross-Sectional Studies (AXIS), Working Group Item Bank (WGIB), and Risk Of Bias In Non-randomised Studies - of Interventions (ROBIN-I; (Page et al., 2018; Sanderson, Tatt, & Higgins, 2007; Viswanathan, Berkman, Dryden, & Hartling, 2013). The ROBIN-I items concerning the randomisation procedure were omitted; an “intervention” was substituted with “exposure”.

### Synthesis methods

Bayesian updating is defined as a procedure of updating prior belief by incorporating new information. Degrees of belief about the probability of an event or an outcome, within a Bayesian prediction model, are represented in the form of a prior distribution (Savage, 1951). Thus, prior distribution is an initial belief about a phenomenon. Likelihood distribution for a belief is the extent to which the hypothesis is likely given the newly observed evidence. Bayesian updating is the process by which the prior changes upon consideration of new evidence. The result of Bayesian updating is a new probability distribution representing the updated belief, which is called posterior probability distribution (Spiegelhalter et al., 2003).

Bayesian meta-analysis was conducted in R^1^ (**Error! Reference source not found.**) following methods outlined in (Spiegelhalter et al., 2003). Bayesian updating was performed to obtain the log Odds Ratio (*log OR*) for the association between physical activity and a barrier or enabler (Roberts et al., 2002). In this review, qualitative evidence was used to elicit the prior distribution, and quantitative evidence was used to elicit the likelihood. Posterior distribution was obtained by updating the qualitative evidence with the quantitative evidence. Detailed statistical analysis is reported in supplement 3. Parameters for prior and likelihood were sampled from a normal distribution: **N**(*µ*, *σ*^2^).

### Prior distribution

The qualitative evidence was synthesised using the Theoretical Domains Framework, TDF (Cane et al., 2012). Three reviewers (AA, BV, AC) independently annotated line-by-line the identified qualitative papers using the TDF. Disagreements were resolved through discussion. Then, based on the findings of the TDF analysis, a prior elicitation task was developed to capture experts’ (n = 6) beliefs about the probability distribution for physical activity conditioned on the constructs identified as relevant in qualitative evidence (i.e. informative prior). A prior elicitation task is described in supplement 3. Six reviewers (AA; LT; BV; NA; AC; AT) completed the expert elicitation task (results are summarised in Figure 1). The reviewers made a judgement on whether the hypothetical HF patient met the recommended levels of physical activity or not. The *log OR* distribution was generated from the results of the expert elicitation task (i.e., prior) using the following parameters for the normal distribution: mean (*µ*) and variance (*σ*^2^). The mean (*µ*) was the *log OR* representing the association between a construct being present in a hypothetical scenario describing a patient and the experts’ judgement that the patient was physically active was calculated. The variance (*σ*^2^) was the sampling variance of the association between a construct being present in a scenario and experts’ judgement that the hypothetical patient in the scenario is likely to be active.

**Figure 1.**
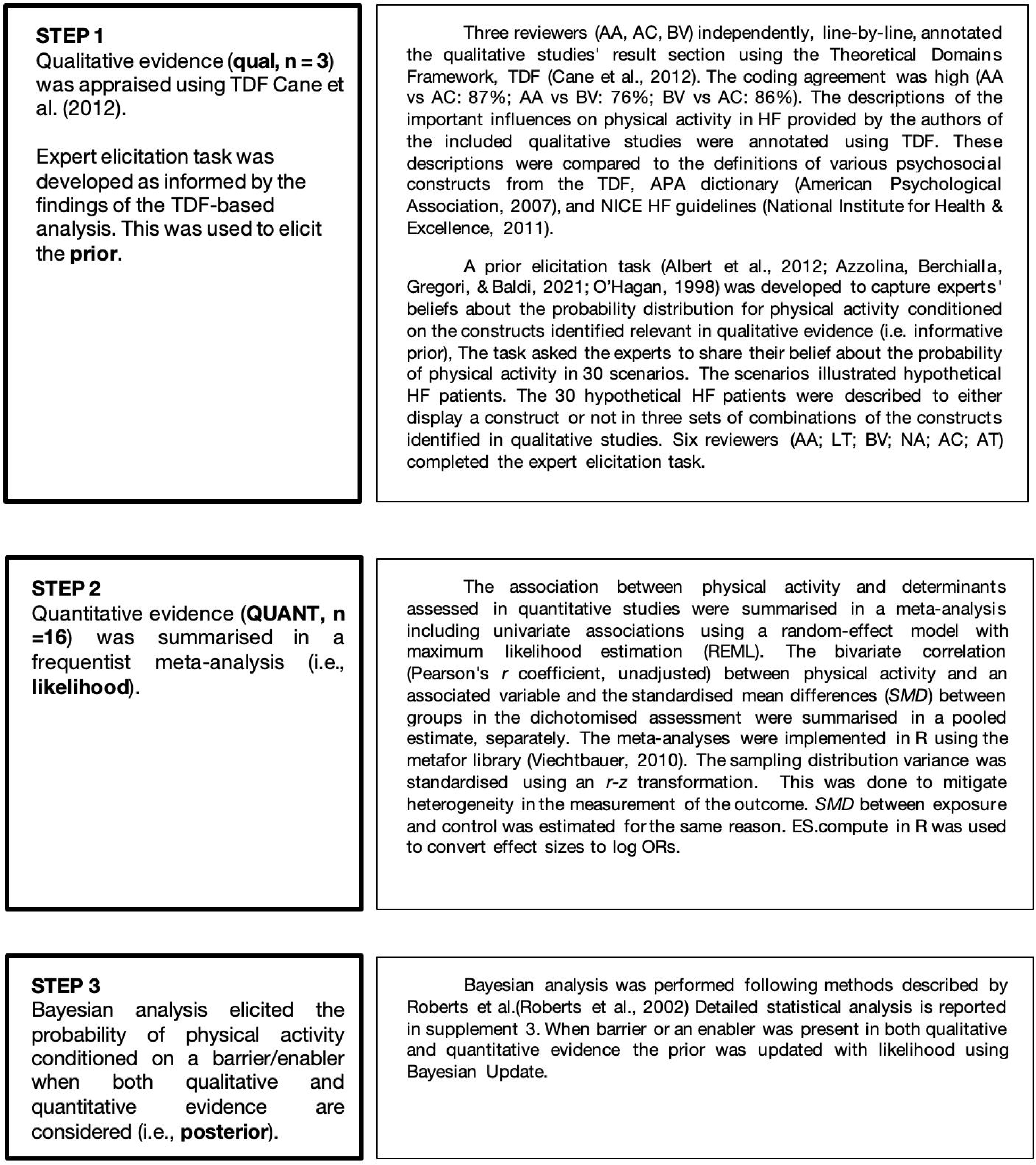
Statistical analysis.

Prior = **N**(*µ*_prior_, *σ*^2^_prior_), where *µ*_prior_ is *log OR* from the expert elicitation task and *σ*^2^_prior_ is sampling variance from the expert elicitation task.

### Likelihood distribution

Effect sizes reported in the individual papers were converted into *log OR using* compute.es library in R (Del Re, 2020). Meta-analysis was performed by pooling individual studies *log OR*s. The likelihood distribution was elicited with the following parameters: the mean (*µ*_likelihood_) was the pooled *log OR* across the included quantitative studies; and the variance (*σ*^2^_likelihood_) was the sampling variance across the included studies.

Likelihood = **N**(*µ*_likelihood_, *σ*^2^_likelihood_), where *µ*_likelihood_ is pooled *log OR* from the meta-analysis including quantitative studies, and *σ*^2^_likelihood_ is sampling variance from the meta-analysis including quantitative studies.

### Posterior distribution

The findings of the prior elicitation task were updated with quantitative evidence concerning each barrier/enabler (i.e., likelihood) using the formula for the Bayesian updating of a normal distribution (p63. Spiegelhalter et al., 2003):

Posterior = **N**(*µ*_posterior_, *σ*^2^_posterior_), where mean is: *µ*_posterior_ = (*µ*_prior_ / *σ*^2^_prior_ + *µ*_likelihood_ / *σ*^2^_likelihood_) / (1/ *σ*^2^_prior_ +1/ *σ*^2^_likelihood_) and variance is: *σ*^2^_posterior_ = 1 / (1/ *σ*^2^_prior_ +1/ *σ*^2^_likelihood_).

### Summary measures

Standardised mean differences (*SMD*) were estimated to describe the impact of exposure on the levels of physical activity as follows: (a) cross-sectional assessment of the differences between the group presenting with a characteristic and the group not presenting with a characteristic (e.g. female = 1; male = 0); (b) pre-post-assessment of physical activity in a cohort study before and after an event of interest (e.g. *SMD* between physical activity outcome before surgery and after surgery); cross-sectional assessment of differences between exercise compliant and non-compliant participants on a range of continuous variables (e.g. *SMD* in self-efficacy between compliers and non-compliers). The r-z transformation was applied in the frequentist meta-analysis of coefficients to mitigate heterogeneity in measurements across studies. The Hartung-Knapp (Sidik-Jonkman) adjustment was made for the evaluation to mitigate small sample size bias (van Aert & Jackson, 2019). compute.es library in R was used to convert effect sizes reported in each included study into *log OR*.

For the Bayesian meta-analysis, the expected value for the *log OR* according to the expert elicitation task, quantitative evidence, and the posterior (qual + QUANT) were calculated. *MAP* and *CrI* as a summary statistic. The dispersion in the probability distribution for log OR associated with each barrier enabler was used to evaluate the level of uncertainty in the evidence in support of that barrier and enabler. The dispersion (i.e., standard deviation, SD) was interpreted relative to the dispersion in the probability distribution for physical activity in the general HF population (i.e., hyperprior) elicited from a large international study (Jaarsma et al., 2013) (see GitHub repository^2^). The SD> 0.70 corresponds to wide dispersion, 0.69-0.21 to medium dispersion, and SD <0.20 corresponds to narrow dispersion.

### Applying findings to intervention development

The identified barriers and enablers in qualitative and quantitative evidence were mapped onto the TDF (Atkins et al., 2017; Cane et al., 2012) and COM-B (Michie et al., 2011). Accordingly, several corresponding behaviour change techniques (BCTs) that are likely to enhance these enablers or reduce the barriers were proposed following the consensus on the link between barriers and enablers and the strategies (Connell et al., 2019).

### Sensitivity analysis

To assess the impact of the qualitative evidence on the findings of this meta-analysis, we performed a sensitivity analysis by excluding the qualitative evidence. In addition, the meta-analysis was stratified by the physical activity outcomes included in the identified studies.

### Certainty assessment

In quantifying the uncertainty in the evidence, the width of the distribution dispersion (SD) was used to estimate the level of uncertainty for each barrier or enabler separately. Wide, medium, and narrow dispersion corresponds to high, moderate, and low uncertainty, respectively.

## Results

### Study selection

The search results are summarised in Figure 1. A total of 9026 titles and abstracts and 80 full-text articles were screened. Nineteen studies cited in supplement 4 (N = 2739) were included in the review. Studies that might appear to meet the inclusion criteria but were excluded, as well as the reasons for exclusion, are reported in supplement 5.

**Figure 1.**
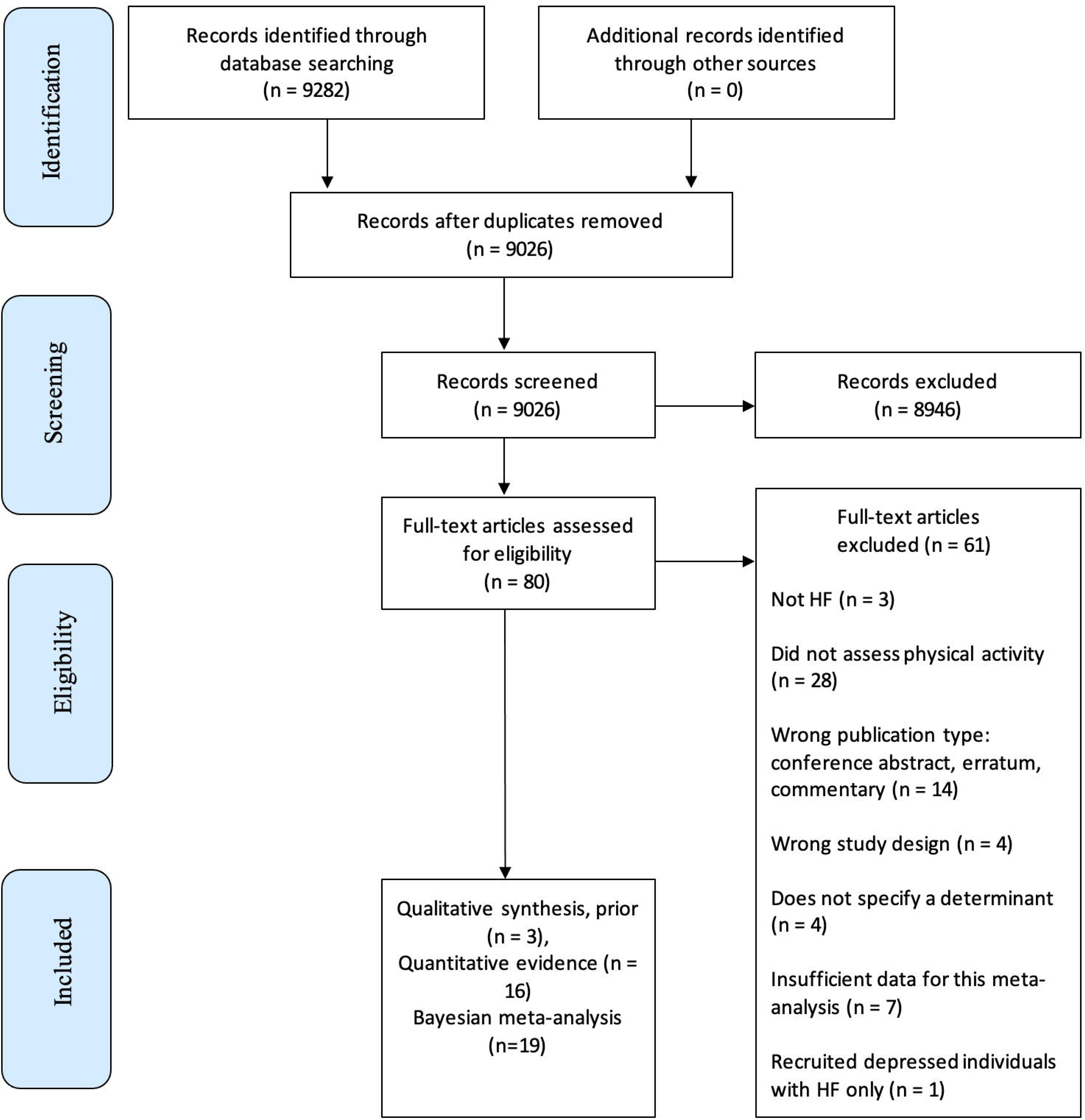
Meta-analysis of barriers and enablers of physical activity in HF: PRISMA diagram

**Figure 2.**
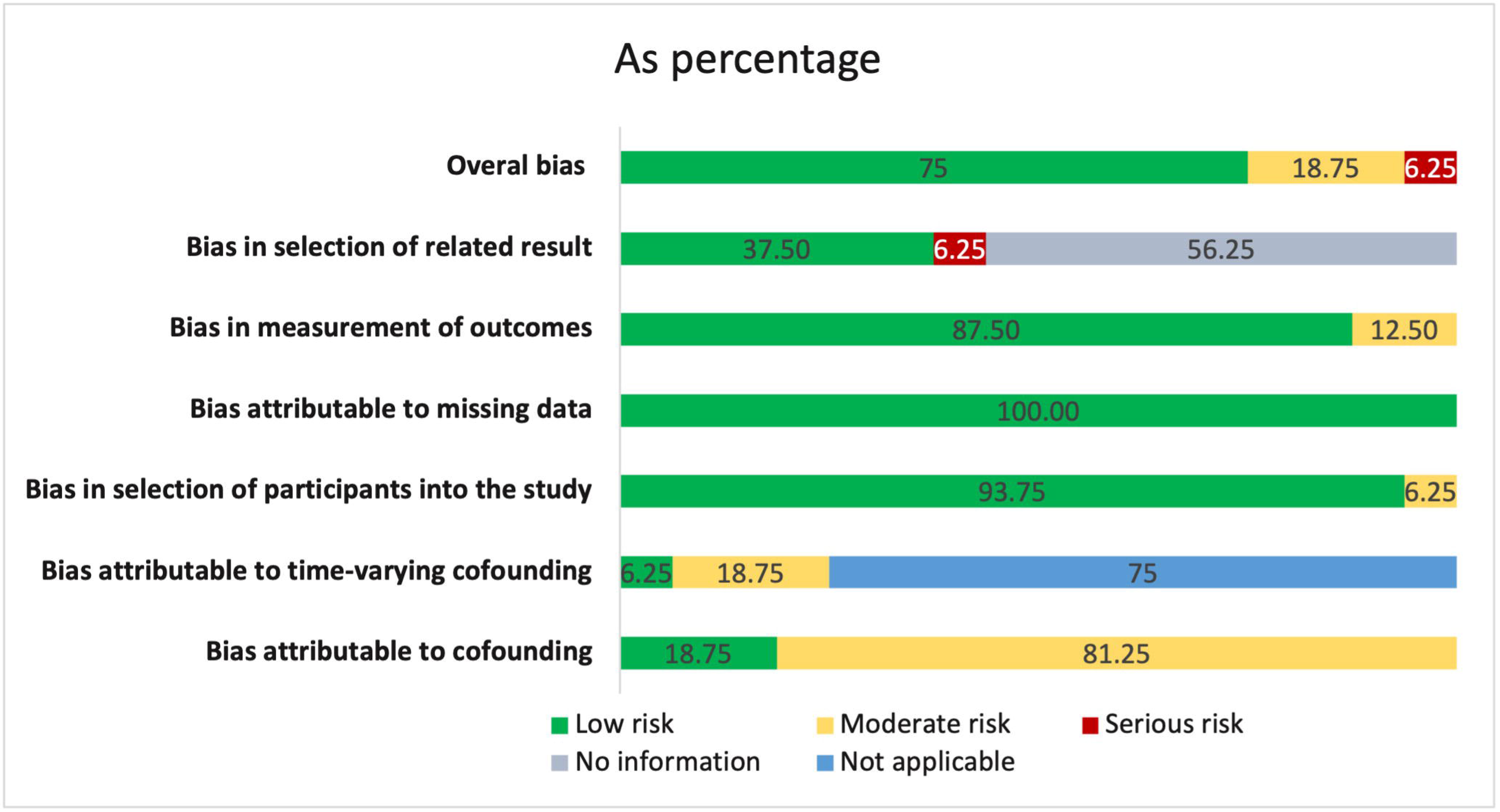
Study-level risk of bias: based on WIB, ROBIN-I, and AXIS items combined into six categories proposed by Page et al., 2018 with an addition of the confounding bias described in ROBIN-I.

### Study characteristics

Studies were conducted in the United States of America (n=8), United Kingdom (n=3), Netherlands (n=2), Sweden (n=2), Australia (n=1), Germany (n=1), Taiwan (n=1), and South Korea (n=1). The majority of the included studies were of a cross-sectional design (n=7, Table 1). The average sample size for quantitative and empirical qualitative studies were 150 and 17, respectively. Physical activity assessment methods and barriers and enablers assessment methods are reported in supplement 6. The mean age of the participants was 63.44 years old (*SD* = 8.39, median = 62.15, *IQR*: [ 59.5; 68]). The Left Ventricular Ejection Fraction (LVEF, %) was moderately low (mean = 34.52%, *SD*=9%). Overall, the majority of samples in the included studies were homogeneous.

**Table 1.**
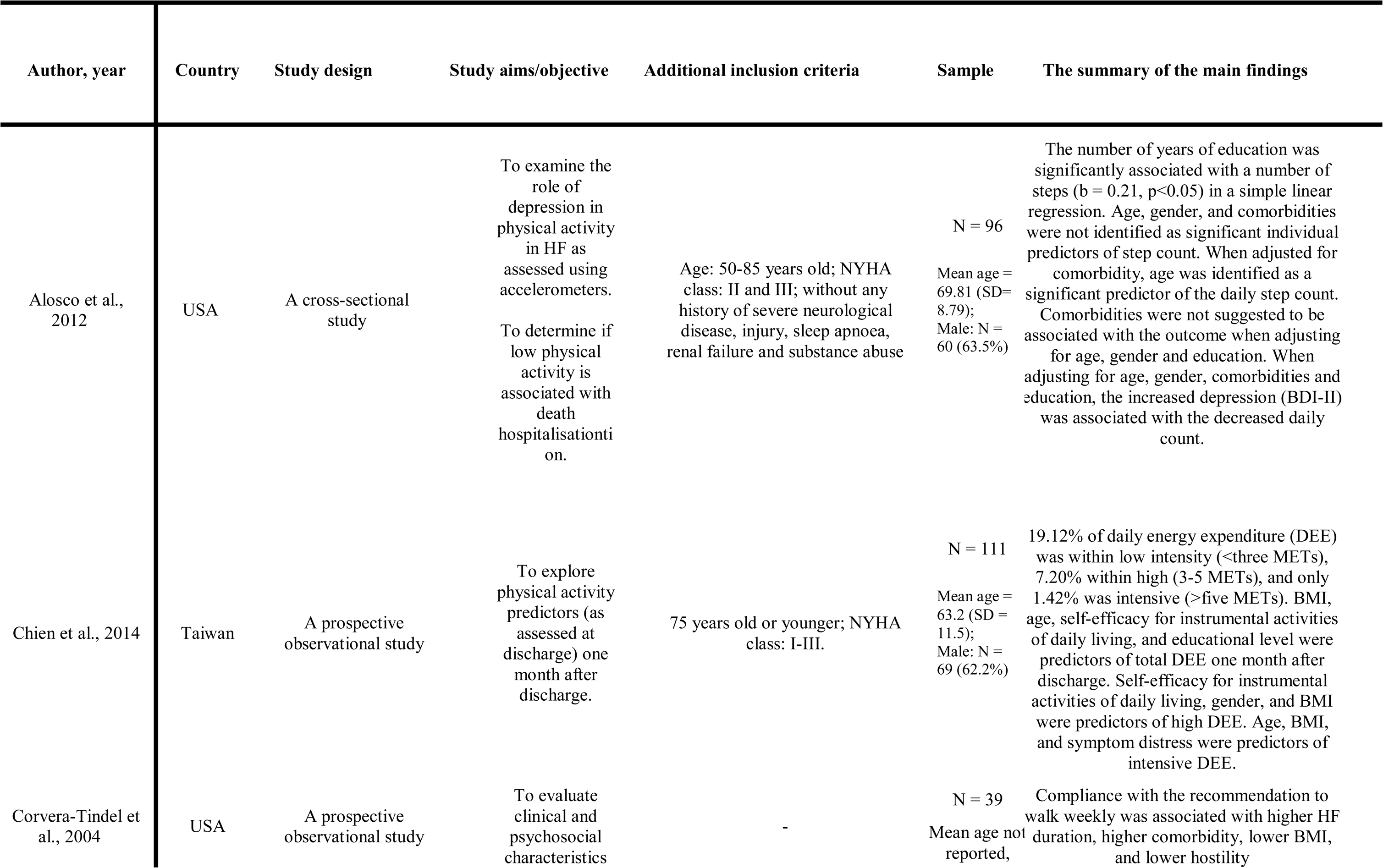
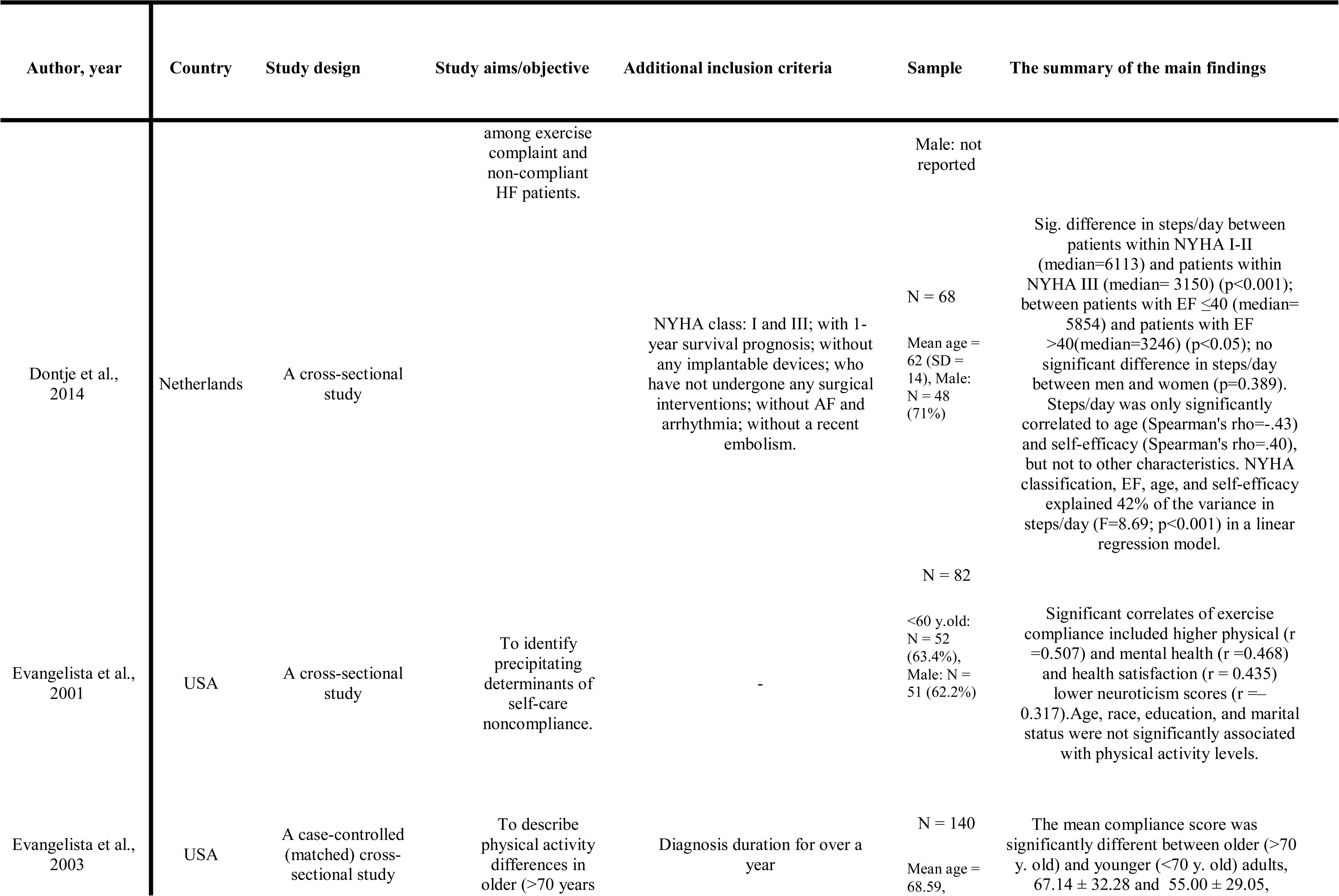
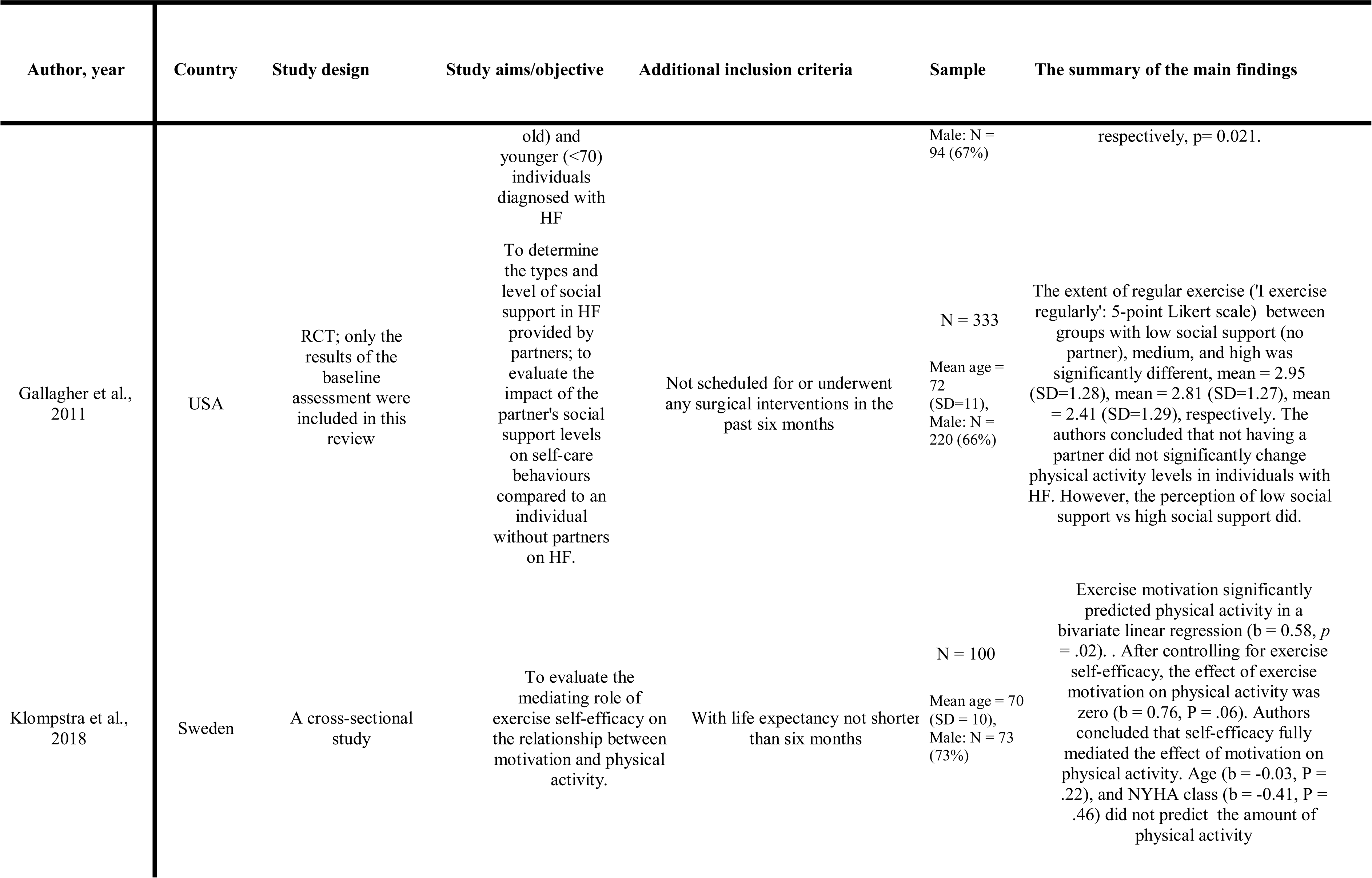
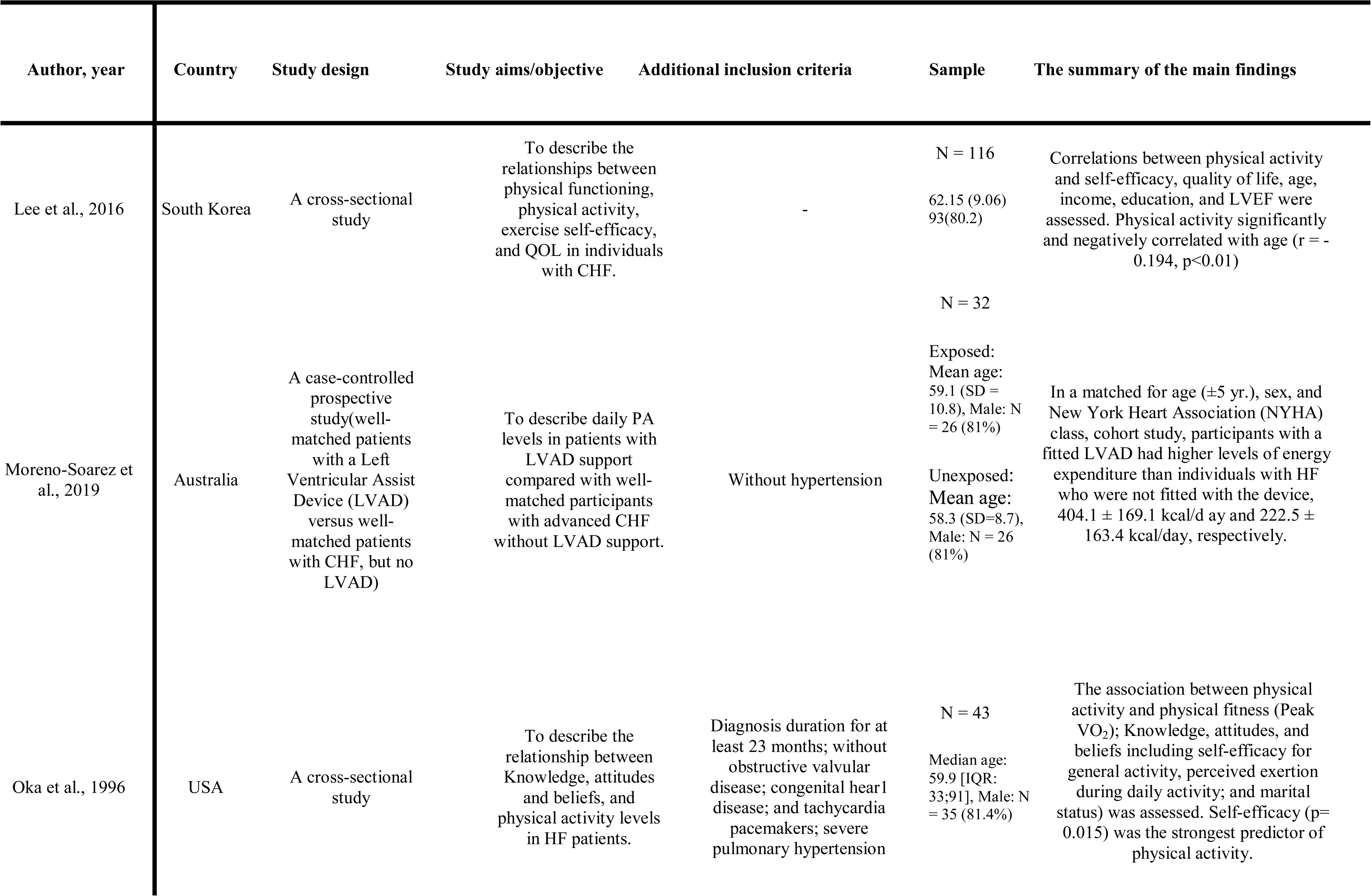
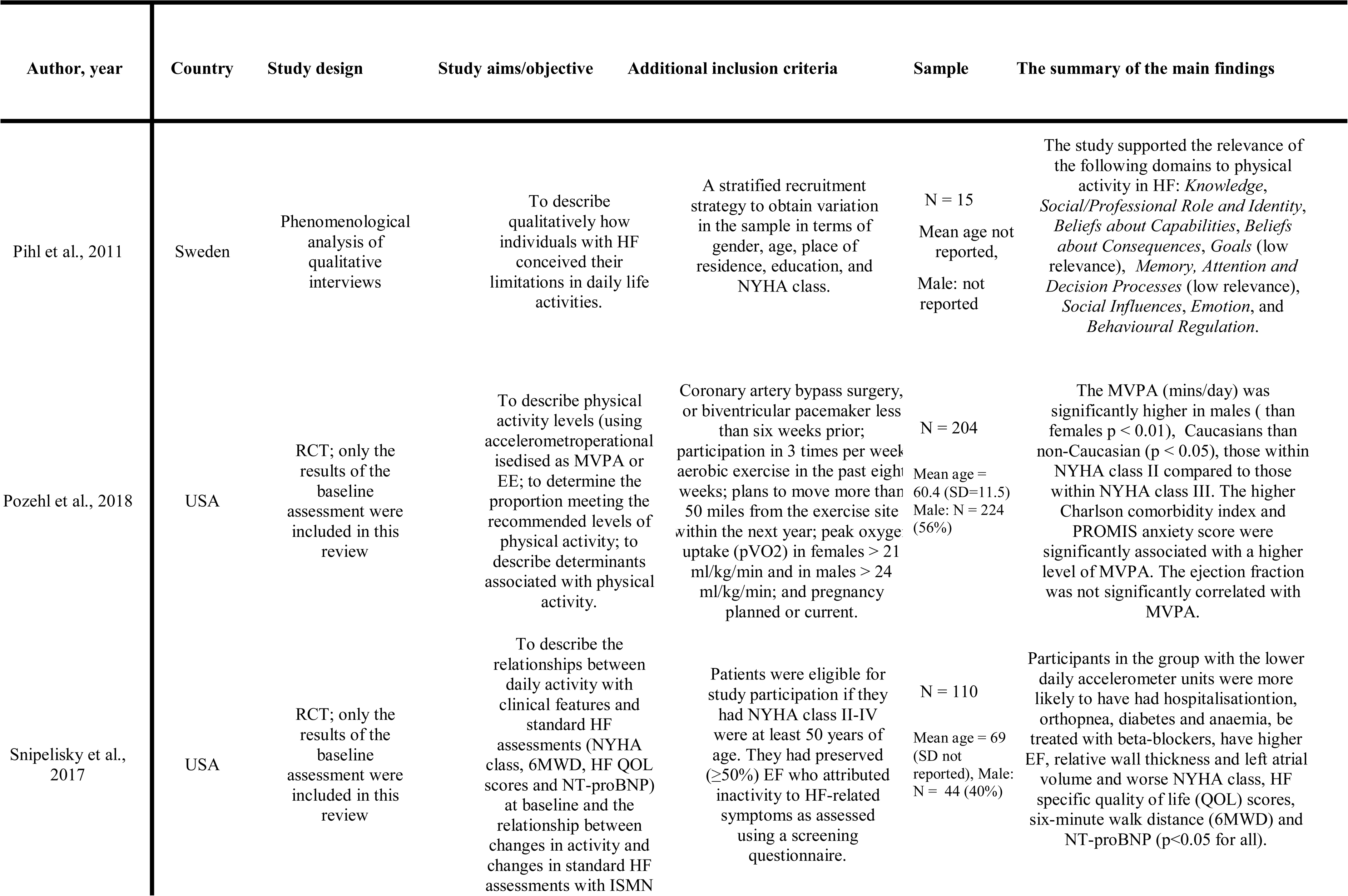
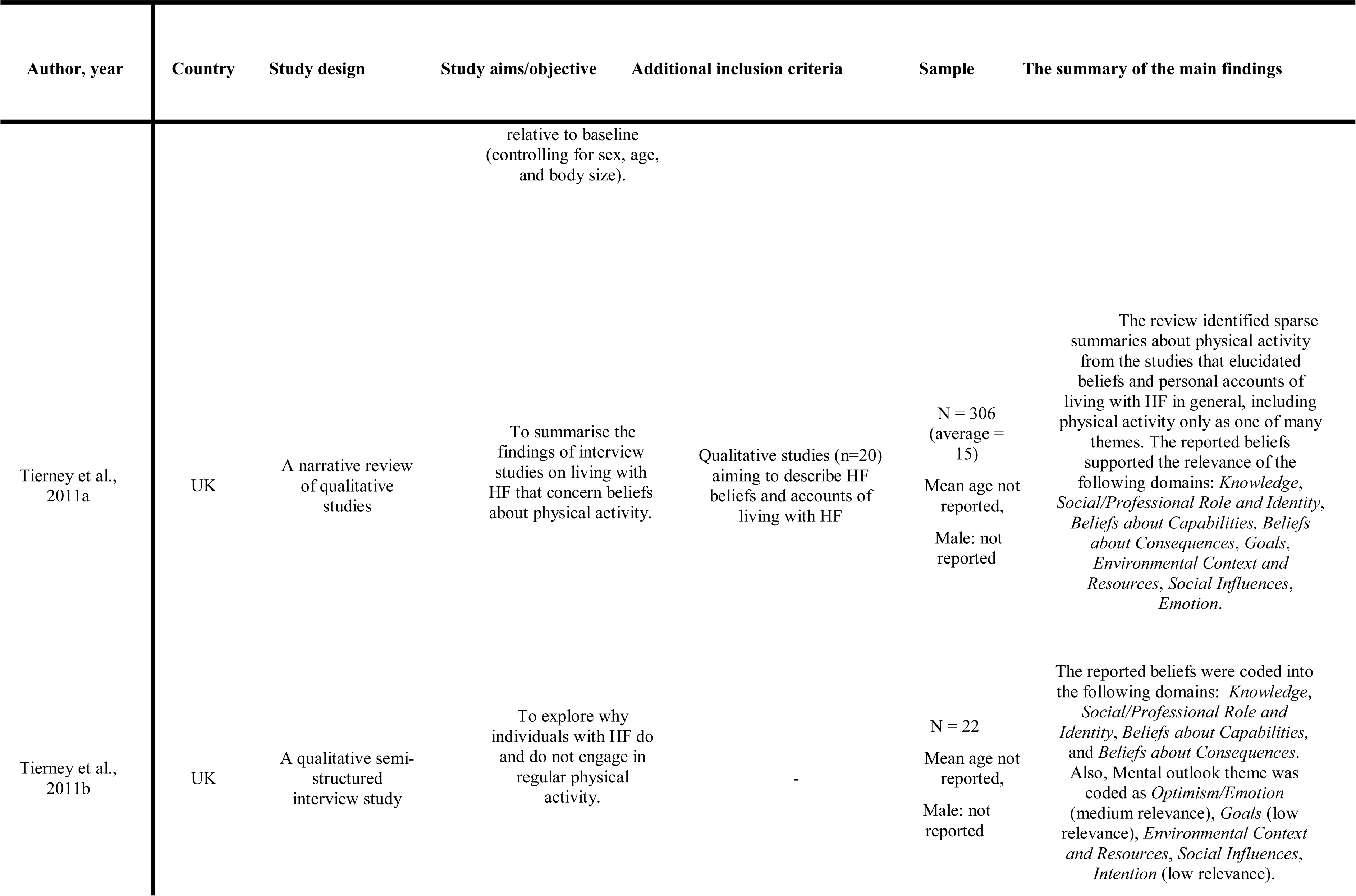
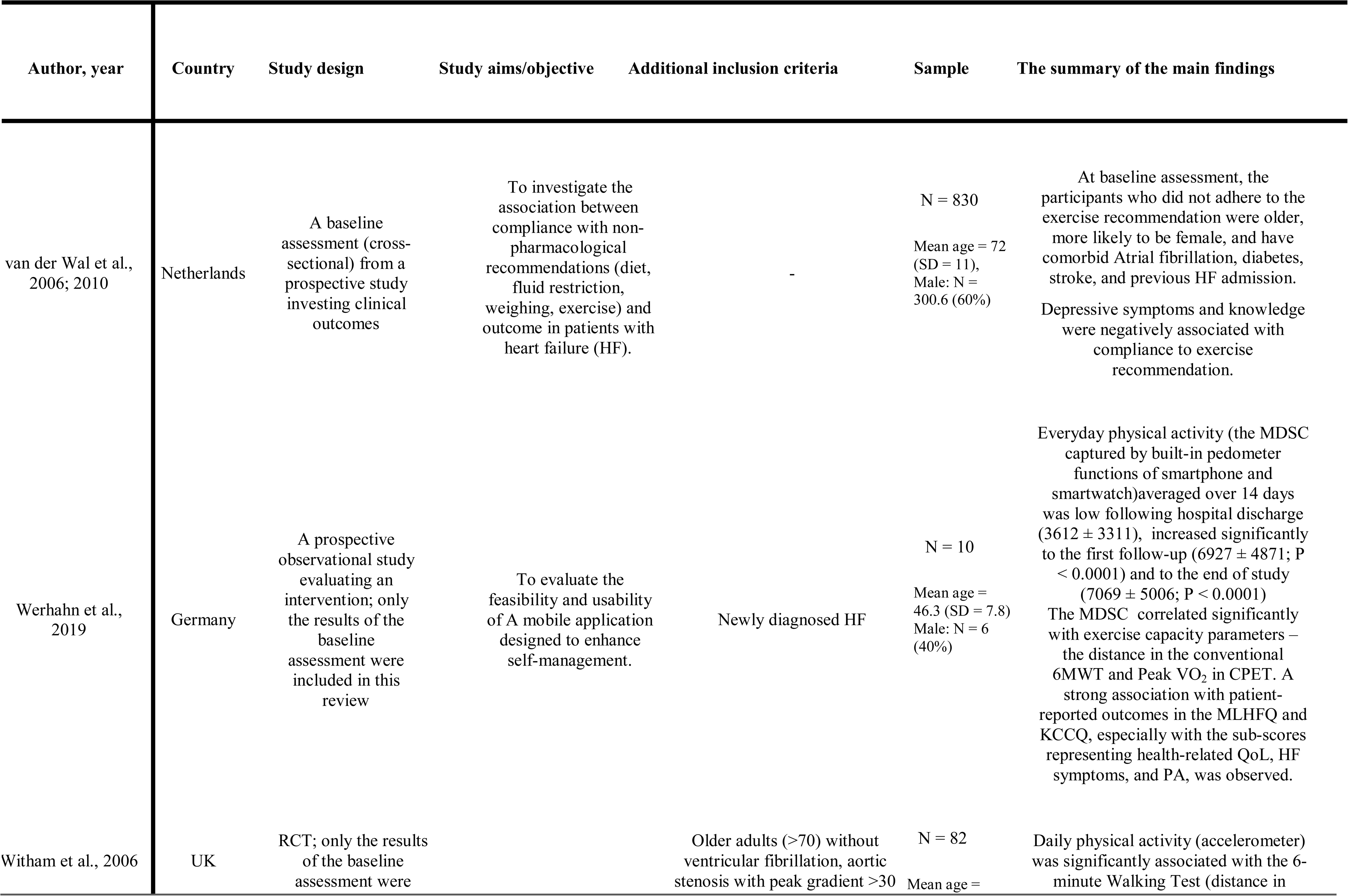
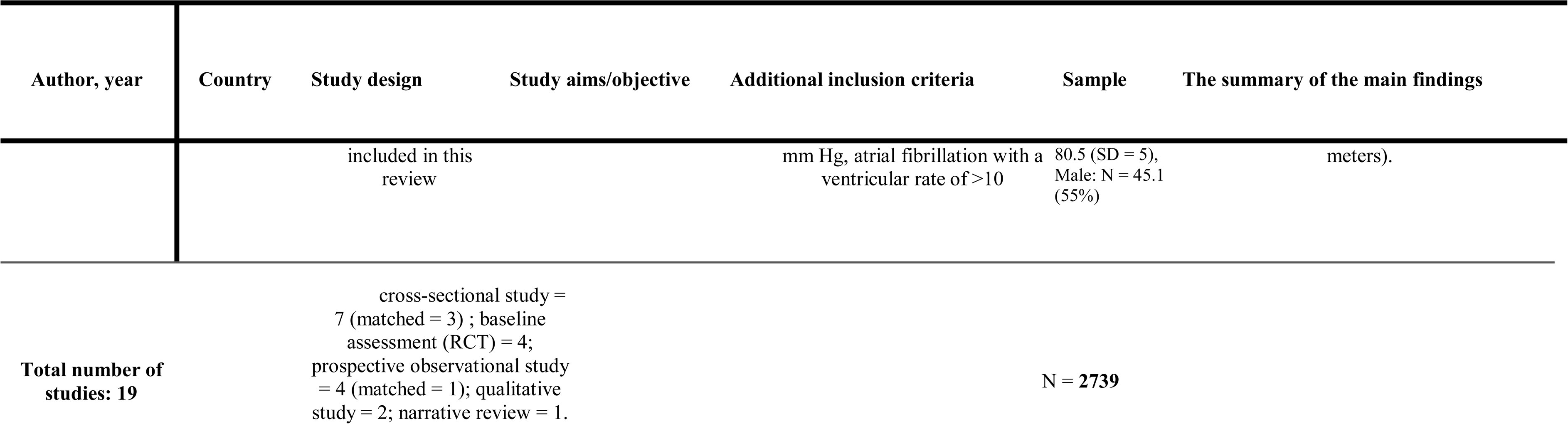
Study characteristics.

### Risk of bias in studies

The risk of bias across the included studies is reported in Error! Reference source not found. The low overall risk of bias was present in 12 (75%) studies, moderate – in three (18.75%) studies (Alosco et al., 2012; Chien et al., 2014; Corvera-Tindel et al., 2004), and serious – in one study (Evangelista et al., 2001). One (6.25%) study was exposed to a serious risk of reporting bias, as only statistically significant results were reported (Evangelista et al., 2001). A total of nine (56.25%) studies did not have a pre-registered protocol, and therefore no information on the bias in the selection of reported results was available. The measurement bias caused by the dichotomisation of the age variable was present in two (12.50%) studies (Evangelista et al., 2001; Evangelista et al., 2003). Participant selection bias was present in one (6.25%) study (Klompstra et al., 2018). Out of four prospective (25%) studies, only one (6.25%) controlled for time-confounding variables by matching participants in exposed and unexposed groups (Moreno-Suarez et al., 2019). Only three (18.75%) studies were exposed to a low risk of bias attributed to confounding: two matched participants (i.e., gender and severity of the disease) when assessing differences in exposed and unexposed groups (Evangelista et al., 2003; Moreno-Soarez et al., 2019), and one measured appropriate confounding variables (Klompstra et al., 2018). The study-level risk of bias assessment is reported in supplement 7.

### Results of synthesis

#### Qualitative evidence

The qualitative evidence synthesis and results are detailed in supplement 3. Overall, the following TDF domains (presented in italics) barriers and enablers influencing physical activity performance by individuals living with HF were found <u>uniquely</u> in qualitative evidence: *Social/Professional Role and Identity*, *Behavioural Regulation*, *Environmental Context and Resources*, *Social Influences*, and *Knowledge* according to three included qualitative studies (Tierney et al., 2011a; Tierney et al., 2011b, Pihl et al., 2011). Coded themes included *’Losing one’s social role in daily life’, which* was annotated as *Social/Professional Role and Identity*. It captured how the loss of participants’ social network and position in society negatively influenced their engagement in physical activity (Pihl et al., 2011). Another theme from the literature, ‘*Need of finding practical solutions in daily life*’ (Pihl et al. 2011), was coded as *Behavioural Regulation* and summarised the need for effective problem solving that enables the integration of physical activity in daily life with ease (supplement 3). One study (Tierney et al., 2011b) identified the relevance of the following TDF domains: *Environmental Context & Resources*, *Social Influences*, *Knowledge* (supplement 3).

The determinants that were reported by both qualitative and quantitative studies were: age, perceived symptoms of HF, functioning, comorbidity, negative attitude, positive physical activity attitude, social support, and self-efficacy. In qualitative studies, the influence of ageing processes was described as ‘*Changing Soma’ (Beliefs about Capabilities)* (Tierney et al., 2011b). Perceived symptoms of HF were described as ‘*Fluctuating health’* (*Beliefs about Consequences)* which impacted physical activity participation (Tierney et al., 2011b). A positive attitude toward physical activity in qualitative studies was described as *’Mental Outlook’ (Belief about Consequences*) (Tierney et al., 2011b), negative attitude in response to physical activity was described as ‘*Negative emotional responses’, (Emotion/Optimism* (Tierney et al., 2011a), social support was described as *’Interpersonal Influences’ (Social Influences)* (Tierney et al., 2011a), and self-efficacy as ‘*Not believing in one’s ability’* (*Beliefs about Capabilities* (Pihl et al., 2011). These qualitative findings informed the expert elicitation task (supplement 3).

#### Bayesian meta-analysis results

The distributions for the log ORs for the association between physical activity and the identified barriers and enablers are described in **Figure 3** and Figure 4. The expected values according to qualitative evidence, quantitative evidence and both are summaries in Table 2.

**Figure 3.**
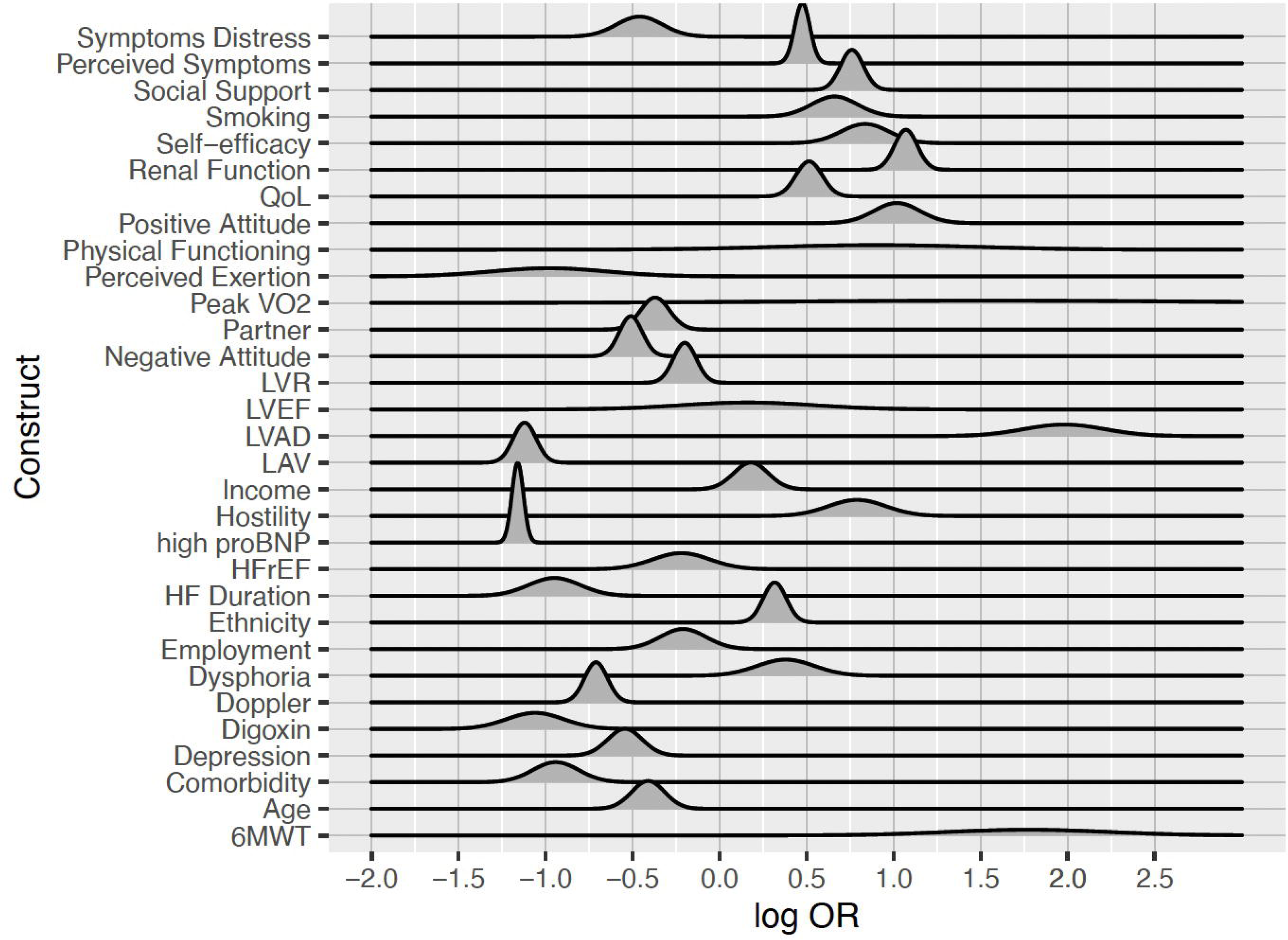
The probability distribution for the expected value of the log OR of physical activity conditioned on identified determinants as suggested by the quantitative evidence (quant), i.e., likelihood.

**Figure 4.**
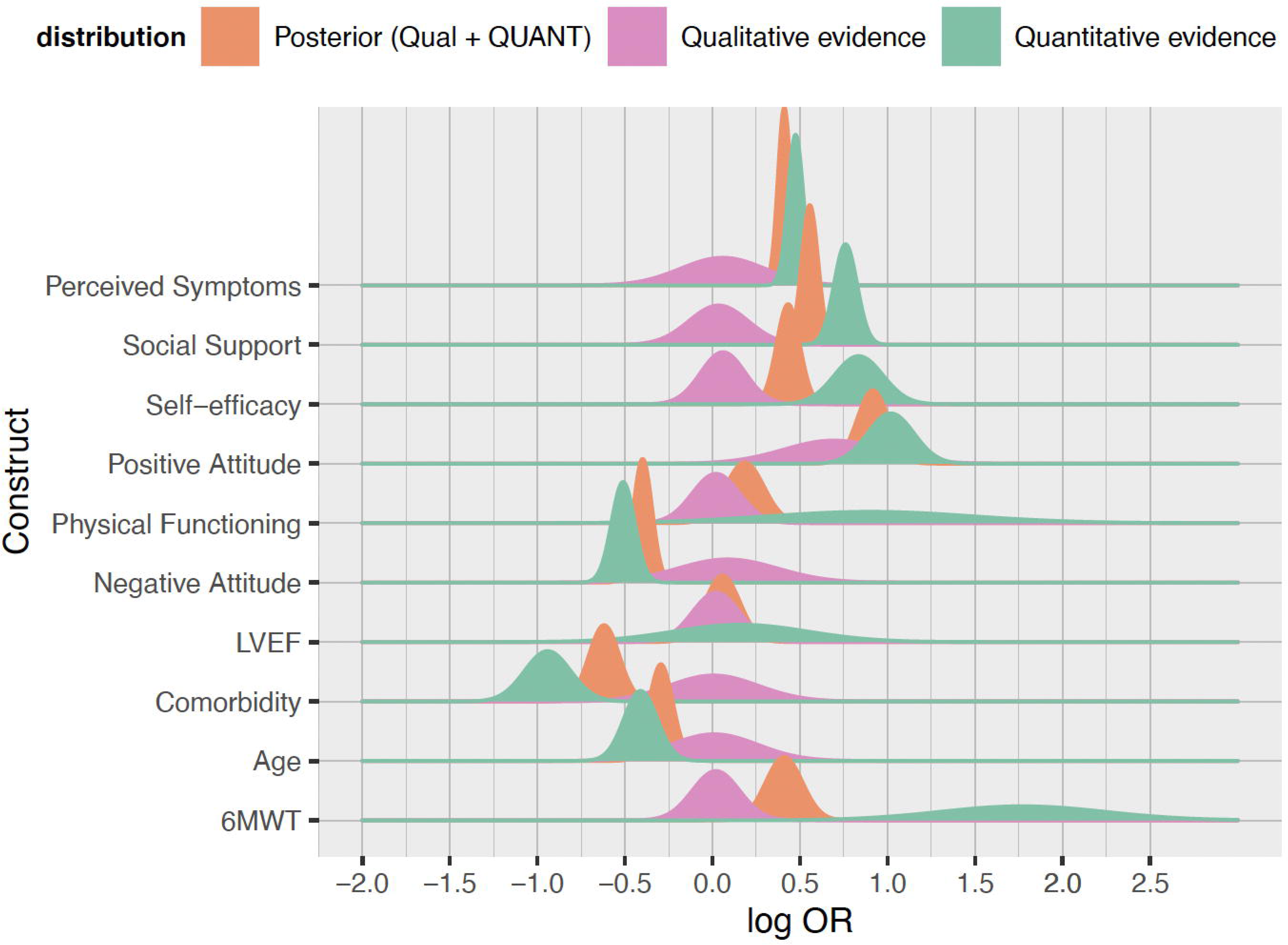
Bayesian updating: the probability distribution for the expected value of the log OR of physical activity conditioned on identified determinants according to qualitative evidence (prior expert elicitation task), quantitative evidence alone (likelihood), and qualitative combined with quantitative evidence (qual + QUANT).

**Table 2.** Summary of the evidence: the expected value for the *log OR* according to the expert elicitation task, quantitative evidence, and the posterior (qual + QUANT) describing the association between physical activity and identified barriers and enablers.

| Construct | Expert elicitation task results (prior) |  |  | Quantitative evidence (likelihood) |  |  | Posterior (qualitative and quantitative evidence) |  |
| --- | --- | --- | --- | --- | --- | --- | --- | --- |
|  | *n qual. studies* | *Expected value (log OR)* | *95% CrI* | *n quant. studies* | *Expected value (log OR)* | *95% CrI* | *MAP (log OR)* | *95% CrI* |
| Age | 3 | 0.01 | [-0.40; 0.42] | 11 | -0.41 | [-0.57; -0.25] | -0.29 | [-0.41; -0.18] |
| Six-minute Walking Test (6MWT) (*Changing Soma* in qualitative evidence) | 3 | 0.02 | [-0.20; 0.24] | 5 | 1.77 | [1.00; 2.54] | 0.41 | [0.24; 0.58] |
| Perceived Symptoms | 2 | 0.06 | [-0.34; 0.45] | 1 | 0.48 | [0.40; 0.55] | 0.41 | [0.35; 0.47] |
| Left Ventricular Ejection Fraction (LVEF), % (*Changing Soma* in qualitative evidence) | 3 | 0.02 | [-0.20; 0.24] | 6 | 0.16 | [-0.47; 0.79] | 0.06 | [-0.11; 0.22] |
| Self-efficacy | 2 | 0.06 | [-0.15; 0.27] | 6 | 0.84 | [0.61; 1.06] | 0.43 | [0.32; 0.54] |
| Social support | 1 | 0.03 | [-0.24; 0.31] | 1 | 0.76 | [0.65; 0.87] | 0.56 | [0.48; 0.63] |
| Comorbidity | 1 | 0.01 | [-0.41; 0.43] | 2 | -0.94 | [-1.16; -0.72] | -0.62 | [-0.76; -0.48] |
| Negative attitude | 1 | 0.09 | [-0.38; 0.56] | 1 | -0.51 | [-0.62; -0.40] | -0.40 | [-0.49; -0.31] |
| Physical functioning | 1 | 0.02 | [-0.20; 0.24] | 4 | 0.90 | [-0.06; 1.86] | 0.18 | [0.01; 0.36] |
| Positive physical activity attitude | 1 | 0.69 | [0.22; 1.16] | 1 | 1.02 | [0.80; 1.23] | 0.92 | [0.77; 1.06] |
| Depression |  | - |  | 6 | -0.54 | [-0.71; -0.38] | - |  |
| Digoxin prescription |  | - |  | 1 | -1.06 | [-1.33; -0.79] | - |  |
| Doppler estimated filling pressure |  | - |  | 1 | -0.71 | [-0.82; -0.60] | - |  |
| Dysphoria |  | - |  | 1 | 0.38 | [0.11; 0.65] | - |  |
| Employment |  | - |  | 2 | -0.21 | [-0.42; 0.01] | - |  |
| Ethnicity (Caucasian vs non-Caucasian) |  | - |  | 2 | 0.32 | [0.21; 0.42] | - |  |
|  | *n qual. studies* | *Expected value (log OR)* | *95% CrI* | *n quant. studies* | *Expected value (log OR)* | *95% CrI* | *MAP (log OR)* | *95% CrI* |
| HF duration |  | - |  | 2 | -0.95 | [-1.19; -0.71] | - |  |
| HFrEF (Yes, vs HFpEF) |  | - |  | 1 | -0.22 | [-0.49; 0.05] | - |  |
| High pro-BNP |  | - |  | 1 | -1.16 | [-1.21; -1.11] | - |  |
| Hostility |  | - |  | 1 | 0.79 | [0.52; 1.06] | - |  |
| Income |  | - |  | 1 | 0.18 | [0.02; 0.34] | - |  |
| Left Atrial Volume index (LAV) |  | - |  | 1 | -1.12 | [-1.23; -1.01] | - |  |
| Left Ventricular Assist Device (LVAD) |  | - |  | 1 | 1.98 | [1.60; 2.36] | - |  |
| Left Ventricular Remodelling (LVR) |  | - |  | 1 | -0.20 | [-0.31; -0.09] | - |  |
| Living with Partner |  | - |  | 2 | -0.37 | [-0.51; -0.24] | - |  |
| Peak VO<sub>2</sub> |  | - |  | 2 | 1.54 | [-0.41; 3.49] | - |  |
| Perceived exertion |  | - |  | 1 | -0.98 | [-1.52; -0.44] | - |  |
| Quality of Life (QoL) |  | - |  | 3 | 0.51 | [0.39; 0.64] | - |  |
| Renal function |  | - |  | 1 | 1.07 | [0.96; 1.18] | - |  |
| Smoking |  | - |  | 1 | 0.66 | [0.44; 0.88] | - |  |
| Symptom distress |  | - |  | 1 | -0.25 | [-0.47; -0.03] | - |  |

**Note.** *OR* – Odds ratio; *CrI* – Credible Interval.

##### Contextual factors

High pro-b-type natriuretic peptide, pro-BNP (*MAP* value for *log OR* = −1.16; 95% *CrI*: [-1.21; - 1.11]) was suggested as a barrier to physical activity with narrow distribution dispersion (*SD* = 0.18). Another contextual barrier with narrow dispersion (*SD* = 0.19) are self-reported symptoms (*MAP* value for *log OR* = 0.48; 95% *CrI*: [0.40; 0.55]). Contextual barriers with moderate uncertainty judged from the distribution dispersion ranging from 0.26 to 0.41 were age (years) (*MAP* value for *log OR* = −0.29; 95% *CrI*: [-0.41; −0.18]), comorbidity measured using Charlson Comorbidity Index (*MAP* value for *log OR* = −0.62; 95% *CrI*: [-0.76; −0.48]), depression measured using HADS-D CES-D, and PROMIS-29 (*MAP* value for *log OR* = −0.54; 95%C *CrI*: [-0.71; −0.38]), digoxin prescription (*MAP* value for *log OR* = −1.06; 95% *CrI*: [- 1.33; −0.79]), high doppler estimated filling pressure (*MAP* value for *log OR* = −0.71; 95%CrI: [-0.82; - 0.60]), HF duration (*MAP* value for *log OR* = −0.95; 95%*CrI*: [-1.19; −0.71]), Left Atrial Volume index, LAV (*MAP* value for *log OR* = −1.12; 95%*CrI*: [-1.23; −1.01]), and living with partner (*MAP* value for *log OR* = - 0.37; 95% *CrI*: [-0.51; −0.24]).

Contextual enablers with medium dispersion (*SD* ranged from 0.26 to 1.09) were physical functioning (measured using MOS SF-36, KCCQ; *MAP* value for *log OR* = 0.18; 95% *CrI*: [0.01; 0.36]), high 6-minute walking test result (6MWT; *MAP* value for *log OR* = 1.77; 95% *CrI*: [1.00; 2.54]), having an implantable device (left ventricular assistant device, LVAD; *MAP* value for *log OR* = 1.98; 95% *CrI*: [1.60; 2.36]), renal function (glomerular filtration rate (ml/min); *MAP* value for *log OR* = 1.07; 95%CrI: [0.96; 1.18]).

##### Modifiable factors

Modifiable barriers were symptom distress (measured using MSAS-SF; *MAP* value for *log OR* = - 0.46; 95%*CrI*: [-0.68; −0.24]), and negative attitude (Negative Attitude Scale; *MAP* value for *log OR* = −0.40; 95% *CrI*: [-0.49; −0.31]), The distribution dispersion of *SD* = 0.36 and 0.26, respectively, indicating a moderate uncertainty in the evidence.

Modifiable enablers were social support (*MAP* value for *log OR* = 0.56; 95% *CrI*: [0.48; 0.63]), self-efficacy (*MAP* value for *log OR* = 0.43; 95% *CrI*: [0.32; 0.54]), positive physical activity attitude (*MAP* value for *log OR* = 0.92; 95% *CrI*: [0.77; 1.06]), distribution dispersion: *SD* = 0.26, 0.37, and 0.36, respectively, indicating moderate uncertainty in the evidence.

#### Sensitivity analysis results

The results of the sensitivity analysis comparing qualitative evidence to quantitative evidence and the results of the analysis combining both are summarised in Figure 4.

Heterogeneous physical activity outcomes were combined in the main meta-analysis. This included accelerometer units (n = 2), duration, mins/day assessed using an accelerometer (n = 1), energy expenditure estimated from accelerometer data (metabolic equivalents, METs; n = 4), self-reported exercise compliance (n = 6), self-reported general physical activity, measured using International Physical Activity Questionnaire, IPAQ (n = 2), self-reported adherence to prescribed exercise self-care behaviour (n = 1), steps per day (n = 3), and one study included both energy expenditure (METs) and duration (mins/day), supplement 6. The results of the analysis evaluating the association between each identified barrier or enabler and the included physical activity outcomes are reported in supplement 8.

The sensitivity analysis results stratified by physical activity outcome assessed in quantitative evidence (likelihood) are reported separately for each barrier and enabler in supplement 8. Sensitivity analysis highlighted the following changes in the evidence compared to the main results. Studies (n = 2) assessing physical activity using an accelerometer did not support depression as a considerable barrier to physical activity. Studies (n = 4) assessing the relationships between energy expenditure (METs) provided evidence with moderate uncertainty regarding perceived symptoms in comparison to the main results suggesting that perceived symptoms are a barrier with low uncertainty in the evidence. Studies (n = 3) that assessed steps per day using an accelerometer suggest considerably high uncertainty in the evidence regarding the barriers (i.e., Pro-BNP) and enablers (i.e., 6MWT, physical functioning, LVEF, Peak VO_2_). The findings of the meta-analysis restricted to the studies assessing self-reported physical activity duration per day, self-reported exercise recommendation compliance, self-reported physical activity (IPAQ), and physical activity as self-care behaviour did not differ from the main results (supplement 8).

### Applying findings to intervention development

Table 3 reports barriers and enablers identified in qualitative evidence that need to be further investigated in quantitative studies (high uncertainty) and barriers and enablers supported in quantitative evidence with low or moderate uncertainty and the behaviour change strategies that may be useful in addressing them.

**Table 3.** Summary of the barriers and enablers suggested by qualitative and quantitative evidence and proposed behaviour change techniques (BCTs).

| Construct | Type of evidence | Distribution SD | Uncertainty in the evidence | COM-B | TDF domain | Mechanisms of Action (MoAs) | Proposed behaviour change techniques (BCTTv1) |
| --- | --- | --- | --- | --- | --- | --- | --- |
| **Contextual factors** |  |  |  |  |  |  |  |
| Perceived Symptoms | Qual + QUANT | 0.19 | narrow/low | Psychological Capability, Physical Capability | - | - | - |
| Age | Qual + QUANT | 0.26 | medium/moderate | Psychological Capability, Physical Capability | - | - | - |
| Comorbidity | Qual + QUANT | 0.29 | medium/moderate | Psychological Capability, Physical Capability | - | - | - |
| 6MWT | Qual + QUANT | 0.32 | medium/moderate | Psychological Capability, Physical Capability | - | - | - |
| Physical Functioning | Qual + QUANT | 0.33 | medium/moderate | Psychological Capability, Physical Capability | - | - | - |
| LVEF | Qual + QUANT | 0.32 | medium/moderate | Psychological Capability, Physical Capability | - | - | - |
| High proBNP | QUANT | 0.18 | narrow/low | - | - | - | - |
| Depression | QUANT | 0.31 | medium/moderate | Automatic Motivation | - | - | - |
| Digoxin | QUANT | 0.41 | medium/moderate | - | - | - | - |
| Doppler | QUANT | 0.26 | medium/moderate | - | - | - | - |
| Dysphoria | QUANT | 0.41 | medium/moderate | - | - | - | - |
| Employment | QUANT | 0.36 | medium/moderate | - | - | - | - |
| Ethnicity | QUANT | 0.26 | medium/moderate | - | - | - | - |
| HF duration | QUANT | 0.38 | medium/moderate | - | - | - | - |
| HFref (Yes) | QUANT | 0.41 | medium/moderate | - | - | - | - |
| Hostility | QUANT | 0.41 | medium/moderate | - | - | - | - |
| Income | QUANT | 0.31 | medium/moderate | - | - | - | - |
| LAV | QUANT | 0.26 | medium/moderate | - | - | - | - |
| LVAD | QUANT | 0.48 | medium/moderate | - | - | - | - |
| LVR | QUANT | 0.26 | medium/moderate | - | - | - | - |
| Living with Partner | QUANT | 0.29 | medium/moderate | - | - | - | - |
| Perceived exertion | QUANT | 0.57 | medium/moderate | - | - | - | - |
| QoL | QUANT | 0.27 | medium/moderate | - | - | - | - |
| Renal function | QUANT | 0.26 | medium/moderate | - | - | - | - |
| Smoking | QUANT | 0.36 | medium/moderate | - | - | - | - |
| PeakVO<sub>2</sub> | QUANT | 1.09 | wide/high | - | - | - | - |
| **Modifiable factors** |  |  |  |  |  |  |  |
| Social support | Qual + QUANT | 0.26 | medium/moderate | Social Opportunity | Social Influences | Social Influences | Social support (unspecified), Social support (emotional), (Social support practical) |
| Negative attitude | Qual + QUANT | 0.26 | medium/moderate | Automatic Motivation | Emotion/Optimism | Emotion | Reduce negative emotions, Information about health consequences, Information about emotional consequences |
| Positive physical activity attitude | Qual + QUANT | 0.36 | medium/moderate | Reflective Motivation | Beliefs about Consequences | Attitude towards the behaviour | Information about consequences, Salience of consequences, Feedback on behaviour, Feedback on the outcome of behaviour, Pros and cons, Emotional consequences, Csensitisation, Anticipated regret, Comparative imagining of future outcomes, Vicarious reinforcement |
| Self-efficacy | Qual + QUANT | 0.37 | medium/moderate | Reflective Motivation | Beliefs about Capabilities | Beliefs about Capabilities | Behavioural practice and Rehearsal, Graded tasks, Social comparison, Focus on past success, Verbal persuasion about capability |
| Symptom distress | QUANT | 0.36 | medium/moderate | Automatic Motivation | Emotion | Emotion | Reduce negative emotions, Information about health consequences, Information about emotional consequences |
| Beliefs about ageing | Qual | - | high | Reflective Motivation | Beliefs about Capabilities | Beliefs about Capabilities | Behavioural practice and Rehearsal, Graded tasks, Social comparison, Focus on past success, Verbal persuasion about capability |
| Social role/self-identity | Qual | - | high | Reflective Motivation | Social/ Professional Role and Identity | Self-image | Identity associated with changed behavior, Reframing, Cognitive dissonance |
| Local | Qual | - | high | Physical Opportunity | Environmental | Environmental | Adding objects to the environment, |
| environment |  |  |  |  | Context and Resources | Context and Resources | Prompts/cues, Avoidance/changing exposure to cues for the behaviour |
| Outcome expectancies | Qual | - | high | Reflective Motivation | Beliefs about Consequences | Beliefs about Consequences | Information about consequences, Salience of consequences, Feedback on behaviour, Feedback on the outcome of behaviour, Pros and cons, Emotional consequences, sensitisation, Anticipated regret, Comparative imagining of future outcomes, Vicarious reinforcement |
| Problem solving | Qual | - | high | Psychological Capability | Behavioural Regulation | Behavioural Regulation | Action planning, Self-monitoring behaviour, Problem solving, Goal setting outcome, Feedback on behaviour, Habit formation |

**Note.** \*The uncertainty in the evidence is judged from the dispersion in the distribution (i.e, SD) relative to the distribution for physical activity in general HF population (SD = 0.14; Jaarsma et al., 2013). The evidence from qualitative studies that was not confirmed in quantitative studies is considered high uncertainty in this review. **QUANT** indicates that the majority of the evidence (n=16) was quantitative, and **qual** indicates that only three studies were qualitative. **SD** – standard deviation; **TDF** – Theoretical Domains Framework (Cane et al., 2012); **COM-B** model (Michie et al. 2011); Mechanisms of action (Connel et al., 2019). **BCTTv1** (Michie et al., 2013).

## Discussion

This review aimed to identify, describe, and compare contextual and modifiable barriers and enablers to physical activity in heart failure (HF) using a Bayesian approach. This work extends the limited research on the modifiable barriers and enablers for physical activity participation by individuals living with HF. Both qualitative and quantitative studies were included in this review and meta-analysis. The contextual barriers supported by the evidence with low uncertainty are high pro-BNP and perceived symptoms. Older age, Left Ventricular Ejection Fraction (LVEF, %), depression, HF duration and living with a partner. Evidence concerning the modifiable barriers: negative attitude and symptom distress; and enablers: social support, positive physical activity attitude, and self-efficacy is moderately uncertain.

This review also aimed to demonstrate the applicability of the Bayesian approach in evidence synthesis informing behaviour change interventions. MRC framework for designing complex interventions (Craig et al., 2008; Skivington et al., 2021) urges researchers to consider evidence from a diverse set of sources when developing interventions. Several methods have been proposed, including the use of mixed-methods studies where one type of evidence (e.g., qualitative) informs the research design of the complimenting study (e.g., quantitative). However, it is less clear how to compare findings from a broad range of studies (qualitative and quantitative) and how to estimate uncertainty in the evidence. The Bayesian approach presents a unique opportunity for research informing complex intervention development by providing a workflow and analysis equipped to combine different evidence and evaluate uncertainty in the evidence. This review synthesised evidence from different sources. The prior elicitation task facilitated this. The results of the expert elicitation task were updated with quantitative evidence. Such Bayesian updating of the probability of physical activity in HF conditioned on each construct summarises both qualitative and quantitative evidence. This approach was first implemented by Dixon-Woods and colleagues (Dixon-Woods, Agarwal, Jones, Young, & Sutton, 2005; Roberts et al., 2002). Dixon-Woods et al. (Dixon-Woods et al., 2005; Roberts et al., 2002) advocated for integrating qualitative research in healthcare decision making because it provides valuable insights and places the patient in the heart of care by bringing their perspective into account. However, Roberts et al. (2002) highlighted the following shortcoming of using Bayesian meta-analysis. Qualitative research should be formally and systematically catalogued before it can be integrated with quantitative findings, which is often not straightforward. Using the TDF (Cane et al., 2012) in the present review, we mitigated this limitation.

### Contextual barriers and enablers

Older age is a barrier to physical activity in HF, as suggested by both qualitative and quantitative evidence. This result further reiterated the finding of a meta-analysis (Amirova et al., 2021) that older adults living with HF need more support to attain higher physical activity levels.

Depression is a considerable barrier, as identified by the quantitative evidence. Depression is a large burden on the HF population. It is associated with poor adherence to pharmaceutical treatment (Goldstein, Gathright, & Garcia, 2017) and is an independent predictor of morbidity (Moudgil & Haddad, 2013). The physiological determinants perpetuating depression in HF include inflammation, blood cell abnormalities, CNS changes and changes in health-protective behaviours (Huffman, Celano, Beach, Motiwala, & Januzzi, 2013). The association between depression, HF, and lack of physical activity is complex. Like any cardiovascular disease, HF is a consequence of low physical activity in clinically depressed individuals (Gold et al., 2020). More research investigating the mechanism via which depression impacts physical activity in HF is needed.

The review findings concerning employment are in accord with a qualitative semi-structured interview study with a non-clinical sample of adults transitioning to retirement, which found that retirement is perceived as providing opportunities to become physically active (McDonald, O’Brien, White, & Sniehotta, 2015). On the other hand, authors also reported that this was not always the case, and an individualised approach may be required (McDonald et al., 2015). Similarly, a national survey of 1550 adults aged 60-69 in England in 2007 reported that work commitments and lack of leisure time were major barriers to physical activity (Chaudhury & Shelton, 2010). Context, social norms surrounding physical activity in older age may impact how physical activity is enacted in older adults who transitioned to retirement (Koeneman, Chorus, Hopman-Rock, & Chinapaw, 2017; McPhee et al., 2016).

The diagnosis of HFrEF and its duration may engender a higher risk of physical *in*activity than the diagnosis of HFpEF. However, the available evidence is uncertain, and more evidence is needed before drawing any definitive conclusions. Non-cardiovascular comorbidities in HF include Diabetes Mellitus (type 2), chronic obstructive pulmonary disease (COPD), and renal dysfunction (Rushton, Satchithananda, Jones, & Kadam, 2015). A frequent comorbid cardiovascular condition is atrial fibrillation (Ling, Kistler, Kalman, Schilling, & Hunter, 2016). These comorbidities increase both morbidity and mortality in HF (Rushton et al., 2015). This review identified that a greater number of comorbidities reduce the physical activity engagement in HF. Another clinical barrier identified by the present review is longer HF duration which is likely to result in deterioration of physical functionating. Overall, it is likely that longer HF diagnosis duration, as well as multimorbidity, contribute to limited physical activity levels.

These contextual factors need to be carefully considered in both future cross-sectional studies and randomised-controlled trials evaluating the mechanism of change. Understanding the contextual determinants influencing behaviour is useful in informing the design of quantitative research studies investigating modifiable determinants influencing physical activity (Rothman, Lash, & Greenland, 2008). Contextual differences (i.e., age, LVEF, and depression) indicate that different approaches to behaviour change interventions for these subgroups that take into account their unique clinical characteristics and align with the European Society of Cardiology (Ponikowski et al., 2016) and NICE (National Institute for Healthcare and Excellence, 2018) guidelines are required. The review encourages the consideration of these patient characteristics in the intervention design and its tailoring. However, contextual understanding does not provide insights into what can and needs to be changed for these demographic and clinical subgroups to engage in physical activity. This urges research on modifiable barriers and enablers to physical activity in HF in these subgroups.

### Modifiable barriers and enablers

Both qualitative and quantitative evidence included in this meta-analysis suggests that perceived symptoms and negative attitude (*Emotion*) are relevant barriers. While social support (*Social Influences*), positive physical activity attitude (*Beliefs about Consequences*) and self-efficacy (*Beliefs about Capabilities*) are suggested as enablers of the behaviour. Another review of qualitative and quantitative studies on barriers and enablers relevant to older adults (65-70 years old) and middle-aged adults (50-64 years old) identified that older adults might rely on social influence, social reinforcement and assistance in managing the change in lifestyle to a greater extent than the middle-aged adults (Spiteri et al., 2019). Older adults require social support in managing HF and daily life.

The following domains were identified uniquely in qualitative research: *Knowledge, Beliefs about Consequences*, *Environmental Context and Resources*, *Behavioural Regulation and Social/Professional Role and Identity*. According to qualitative evidence alone, individuals living with HF are driven by the motivation to achieve the desired outcome, such as reduced symptoms and improved health (*Beliefs about Consequences*). According to qualitative research, the local environment that encouraged physical activity (e.g., parks; *Environmental Context and Resources*) was fundamental for physical activity enactment. The need to find practical solutions to overcome limitations in physical activity (i.e., problem solving; *Behavioural Regulation*) played a crucial role in physical activity, according to the qualitative studies included in this review. While the change in perceived social role, described as a loss of social network and position in society brought about as a result of HF, had negative implications for physical activity (*Social/Professional Role and Identity*). However, these were not followed up with a quantitative study to confirm their relevance in a larger sample. This meta-analysis suggests exploring and confirming the role of these barriers and enablers in quantitative research.

### Study-level limitations

Currently, there is no gold standard risk of bias assessment for observational studies (Page et al., 2018). Therefore, this review included categories of sources of bias traditionally proposed for assessing study-level bias. These include confounding bias, selection bias, measurement bias, missing data bias, and reporting bias (Page et al., 2018). These collectively formed the criteria for evaluating the risk of bias across the included studies. Overall, the majority of the studies (75%) were exposed to a low risk of bias. The major source of bias in the included studies is confounding, as observational studies included in the review (81.25%) did not control for confounding effects when assessing correlates of physical activity.

### Strength and limitations of this review

We have adhered to the criteria for Bayesian research in conducting this review, supplement 9 (Depaoli et al., 2017). However, there are a few limitations. First, this meta-analysis offered claims about the association, not causality. Second, the prior was elicited using an expert elicitation task with a limited panel of experts. Health psychology researchers appraised qualitative evidence. They then completed a task designed to elicit a prior probability for physical activity conditioned on the constructs identified in the included qualitative studies. While this is an established technique for formalinising an informative prior, it is by definition subjective and thus depends strongly on the members of the expert panel (Albert et al., 2012). In this case, the panel was limited to health psychologists. It would have been beneficial to include other stakeholders, such as HF nurses or cardiologists. Third, the search was performed in January 2020 before the COVID-19 pandemic and has not been updated because the studies conducted since investigated the impact of COVID-19 and the lockdown on physical activity in HF. Such a global change posits an incomparable barrier to physical activity and is difficult to assess alongside general barriers and enablers summarised in this review. Heterogeneous physical activity outcomes were combined in the main meta-analysis. The qualitative evidence described physical activity in the general sense, while quantitative evidence included well-defined heterogeneous physical activity outcomes. Due to this limitation inherited from the qualitative data, in the main Bayesian meta-analysis, we combined heterogeneous physical activity outcomes. In a sensitivity analysis, we assessed the impact of this on the findings, where we stratified the analysis of quantitative evidence by physical activity outcome (supplement 8). Physical activity outcomes included accelerometer units (*n* = 2), duration, mins/day assessed using an accelerometer (*n* = 1), energy expenditure estimated from accelerometer data (METs; *n* = 4), self-reported exercise compliance (*n* = 6), self-reported general physical activity, IPAQ (*n* = 2), self-reported adherence to prescribed exercise self-care behaviour (*n* = 1), steps per day (*n* = 3), and one study included both energy expenditure (METs) and duration (mins/day). The sensitivity analysis results revealed that depression is not a considerable barrier to physical activity assessed using an accelerometer, suggesting that studies assessing physical activity using self-reports may overestimate the impact of depression on actual physical activity levels. The evidence concerning perceived symptoms is moderately uncertain when only energy expenditure (METs) is included in the analysis. While steps per day are not associated with pro-BNP and enablers such as 6MWT, physical functioning, LVEF, and Peak VO_2._ The latter may have resulted from the smaller number of studies being included in the stratified analysis. The findings concerning other barriers and enablers did not change once the analysis was stratified by physical activity outcome. Physical activity behaviour may be more homogeneous in HF than in the general population due to its low levels and physical limitations.

### Recommendations for future research and clinical practice

Older adults (>70 years old) living with HF are at risk of low physical activity levels. It is important to explore beliefs about physical activity that are associated with older age. Research informing the development of interventions for this subgroup of the population is needed. The quantitative evidence suggests that physical activity levels are reduced in the presence of depression (*Emotion*). A better understanding of the mechanism through which depression impacts physical activity in HF and how it can be mitigated is needed. The quantitative evidence on physical activity conditioned on other clinical, demographic, and psychosocial barriers and enablers is uncertain. Research investigating a broad range of clinical, demographic, and psychosocial barriers and enablers to physical activity in HF is warranted. In addition, identified studies did not explore the mechanism underlying physical activity enactment, including how the barriers and enablers interact, which should be further explored in future research.

Finally, tentative suggestions are made for what a future physical activity intervention needs to include Overall, the review findings indicate that to reduce the barriers and enhance the enablers, a behaviour change intervention containing the following BCTs are needed: identity associated with target behaviour, prompt/cues and adding objects to the environment, behavioural practice/rehearsal, and graded tasks. A previous meta-analysis of randomised controlled trials also suggested that these strategies are associated with the efficacy of interventions (Amirova et al., 2021). In addition, the qualitative evidence included in this review suggests that addressing the change in the social identity as a result of acquired HF diagnosis and the perceived appropriateness of physical activity in this context need to be addressed. *Social Influences, Beliefs about Consequences, Behavioural Regulation, and Emotion* via BCTs such as social support, information about health and emotional consequences of the behaviour, problem solving, and reducing negative attitude (*Emotion*), may be effective in increasing physical activity in HF, according to the present review. However, the latter suggestions need to be considered with caution, considering the high uncertainty in the evidence.

## Conclusion

The identified contextual barriers and enablers to physical activity in HF need to be carefully considered when designing interventions and randomised controlled trials evaluating interventions. There is moderate evidence in support of the modifiable barriers – symptom distress (*Emotion*) and negative attitude (*Emotion*) – and modifiable enablers – social support (*Social Influences*), self-efficacy (*Beliefs about Capabilities*), and positive attitude towards physical activity (*Beliefs about Consequences*). Interventions targeting these barriers and enablers warrant further investigation.

The Bayesian approach in this review enabled comparative predictions about barriers and enablers, helped elicit the extent of uncertainty in the evidence and enabled the combination of qualitative and quantitative evidence in a single synthesis. Thus, the present review supports the usefulness of the Bayesian approach to evidence synthesis concerning barriers and enablers to behaviour and in the development of behaviour change interventions.

## Data Availability

Protocol:
The review's protocol was registered on PROSPERO: CRD42021232048.
Availability of data, code and other materials:
https://github.com/AliyaAM/bayesian_meta_analysis.

## Protocol

The review’s protocol was registered on PROSPERO: CRD42021232048

## Funding

This research was supported by a PhD studentship awarded to AA by City University of London.

**Availability of data, code, and other materials:** https://github.com/AliyaAM/bayesian_meta_analysis

## Authors contribution

AA and LT searched electronic databases, performed screening of titles and abstracts and full texts, and carried out the risk of bias evaluation. The qualitative TDF-based analysis was carried out by AA, BV, and AC. The expert elicitation task was completed by AA, LT, BV, NA, AC, and TF. The Bayesian meta-analysis was carried out by AA. All authors contributed to the interoperation of the results and the write-up of the manuscripts.

## Supplements

1. Detailed Inclusion criteria. The scope of the review: pi(e)cos and spider
2. Search strategy.
3. Statistical analysis.
4. The citations for the included studies.
5. Studies that appear to meet the inclusion but were excluded (with the reasons for exclusion).
6. Physical activity assessment across studies.
7. Individual studies risk of bias.
8. The results of the sensitivity analysis stratified by physical activity outcome.
9. Checklist: criteria for evaluating studies employing Bayesian statistics (Depaoli, Rus, Clifton, van de Schoot, & Tiemensma, 2017).
10. PRISMA checklist: https://prisma.shinyapps.io/checklist/.

## Footnotes

1 GitHub repository: https://github.com/AliyaAM/bayesian_meta_analysis.

2 GitHub repository: https://github.com/AliyaAM/bayesian_meta_analysis/blob/main/results_ordered_by_uncertainty.csv https://github.com/AliyaAM/bayesian_meta_analysis/blob/main/dispersion_uncertainty.R

